# Development and Validation of the Healthcare Worker Culture of Support Scale: Preliminary Evidence of Validity and Reliability

**DOI:** 10.64898/2026.02.12.26346214

**Authors:** Monirah Albathi, Alden Gross, Christine Weston, Cheryl Connors, Mansoor Malik, Albert W. Wu

## Abstract

**Objective:** To develop and conduct preliminary testing of the reliability and validity of the Healthcare Worker (HCW) Culture of Support Scale (COS), intended to assess health worker perceptions of institutional support resources and organizational culture related to their well-being.

**Methods:** A cross-sectional survey was conducted with 533 HCWs from ambulatory clinic and rural hospital settings. The survey included validated measures and newly developed items. Exploratory and confirmatory factor analyses (EFA/CFA) were employed to determine the factor structure. Internal consistency and construct validity were assessed using Cronbach’s α and correlation with mental health outcomes.

**Results:** The COS demonstrated a robust three-factor structure: 1) Organizational Support (α = 0.83), 2) Access to Peer Support (α = 0.92), and 3) Availability of Support (α = 0.97), accounting for 84.9% of variance. Cronbach’s alpha for the overall scale was 0.94. CFA confirmed excellent model fit (RMSEA =.049, CFI =.992). Higher COS scores correlated with lower burnout (r = -.47, p <.001) and anxiety (r = -.35, p <.001), and greater resilience (r =.30, p <.001).

**Conclusion:** Preliminary evidence suggests that the COS is a reliable and valid measure of HCWs’ perceptions of organizational support for worker well-being. This scale and the three subscales can provide healthcare institutions with a way to evaluate organizational initiatives to enhance worker well-being and workforce resilience. Further testing is recommended in diverse settings.

## Introduction

The well-being of healthcare workers (HCWs) is a crucial public healthcare concern, directly impacting patient care quality and service delivery (Olagunju et al., 2021; Selamu et al., 2017). Burnout among frontline HCWs is a persistent challenge for health systems, contributing to staff turnover, workforce shortages, and increased risk of patient injuries (Denning et al., 2021; Heath et al., 2020), especially in the context of stressful events such as global pandemics, job actions, and economic uncertainty (Lotta et al., 2021; O’Brien et al., 2022). High levels of anxiety and low morale during the COVID-19 pandemic brought increased attention to the resilience and psychological well-being of frontline HCWs (Rapisarda et al., 2020). HCW burnout has continued as a significant challenge in the face of chronic understaffing, heavy workload and financial pressures. A 2025 nursing survey found that 65% of nurses report high levels of stress and burnout (Galoustian et al., 2025).

Healthcare organizations are increasingly seeking ways to monitor worker well-being and pursue interventions that support workers (Heath et al., 2020). However, there is a potential disconnect between a focus by institutions on physical health metrics and the priority placed by healthcare workers on psychological well-being (Whitcombe et al., 2016). This suggests the need for well-communicated initiatives that align with employee needs to foster trust and ensure program effectiveness (Eisenberger et al., 1986).

Organizational support of HCWs plays an important role in mitigating burnout. Perceived organizational support, including adequate resources, managerial responsiveness, and mental health initiatives, can significantly improve worker well-being and reduce levels of burnout (Whitcombe et al., 2016; Sexton 2021). Conversely, inadequate support exacerbates burnout, leading to decreased productivity and higher turnover rates (West et al., 2018). Therefore, fostering a supportive organizational culture is essential to sustain healthcare workers’ well-being.

Organizational support is critical for HCWs who become “second victims,” (HCWs emotionally traumatized after adverse patient events or medical errors) who often experience guilt, fear, and impaired professional performance (Wu, 2000; Scott et al., 2009). Recognizing this, regulatory bodies like the Joint Commission have advocated for institutional support systems (Edrees et al., 2016; Joint Commission, 2018; Joint Commission, 2020). The Second Victim Experience and Support Tool (SVEST; Burlison et al., 2017), and the WITHSTAND-PSY questionnaire (Busch, 2023) were developed to assess post-event distress. However, they do not measure worker awareness of available institutional support resources or perceptions of the organizational culture of support. Currently there is no psychometric instrument available to measure organizational support for HCW well-being. The 10-item Perceived Organizational Support Questionnaire (Eisenberger, 2008) measures perceived support but does not address the provision and accessibility of support and is not specific to healthcare. This gap highlights the need for a validated instrument to evaluate HCWs’ awareness of institutional support programs and their effectiveness in fostering a culture that promotes well-being and destigmatizes help-seeking after adverse events. The aim of this study was to develop a self-report instrument to measure HCW awareness and attitudes regarding institutional support programs and evaluate the psychometric properties of the new instrument.

## Methods

This was a cross-sectional analysis of survey data from healthcare workers in ambulatory practices and rural hospitals. The current study was a sub-study of a US Health Resources and Services Administration (HRSA)-funded initiative to implement and evaluate the Resilience in Stressful Events (RISE) peer support program in settings outside of academic medical centers. The RISE program, originally developed at Johns Hopkins Hospital (Edrees et al., 2016), provides confidential peer support to healthcare workers following stressful work-related events and has shown effectiveness in reducing burnout while improving resilience and perceptions of organizational support (Moran et al., 2020, Connors et al., 2019; Connors et al., 2024).

Data for this study were collected at baseline from 46 ambulatory care clinics and 2 rural hospitals prior to their implementing RISE (Sites 1, 2, and 3, respectively). Participants completed an online survey via Qualtrics (Provo, UT) between January-February 2023, receiving $20 compensation. Eligible participants included all patient-facing staff at participating institutions. The survey included components to measure the psychological well-being of workers, and worker perceptions of the culture of well-being and support available to them. The well-being of individual workers was assessed using validated measures of anxiety, burnout, and personal resilience (The Generalized Anxiety Disorder-7 (GAD-7; Spitzer et al., 2006) for anxiety symptoms; the Maslach Burnout Inventory (MBI-2, Maslach & Jackson, 1986; West et al., 2012; Li-Sauerwine et al., 2020) for burnout levels; and the abbreviated Connor-Davidson Resilience Scale (CD-RISC2; Vaishnavi et al., 2007) for personal resilience.

A workplace culture of well-being has been described as one that has been designed to support employee well-being (Gunther et al., 2019). This can be evaluated by worker reports or ratings of awareness of resources to support them, and perceptions of organizational commitment to providing support. This analysis employed the baseline survey data to develop and perform initial testing of scales designed to measure health worker attitudes and perceptions of the culture of emotional support at their institution.

### Survey Development

The multidisciplinary project team developed the survey instrument, which also included demographic items (age, gender identity, race, job title, and employment duration) and assessed multiple dimensions: psychological distress, support systems (coworker, supervisor, institutional, and non-work-related), and professional self-efficacy.

The survey comprised both newly developed and validated measures. We included 5 items from the National Institute for Occupational Safety and Health (NIOSH) Worker Well-Being Questionnaire assessing job satisfaction, employees’ perceptions of their ability to count on their supervisor and coworkers for support, trust in the management of their organization, and their perception of their organization’s commitment to employee health and well-being (rated on a 4-point scale from 1=not at all satisfied to 4=very satisfied).

We created new questions where there were not suitable validated items or scales. These included three questions pertaining to supervisor and organizational support; three questions pertaining to the availability of confidential peer support resources within their organization, and five questions pertaining to the availability of someone in their workplace to provide support in the event of a stressful work-related event (rated on 5-point Likert scale 1 = “strongly disagree” to 5 = “strongly agree”). This 11-item Healthcare Worker Culture of Support Scale (COS), can be found in Appendix 1.

### Analytic Approach

The COS was assessed through a rigorous psychometric analysis. Initial exploratory factor analysis (EFA) excluded items assessing overall well-being, resilience, and job satisfaction (Items 1-3) due to potential confounding from external factors, focusing instead on organizational and peer support constructs (Items 4-11). Principal component analysis with oblique rotation (Costello & Osborne, 2005) and Bayesian Information Criterion (BIC) determined the optimal factor structure, followed by confirmatory factor analysis (CFA) assessing model fit, comparative fit index (CFI), and root mean square error of approximation (RMSEA) (Hu & Bentler, 1999). Internal consistency was established using Cronbach’s α and inter-item correlations. We examined scale validity through Pearson/Spearman correlations with established measures (GAD-7: Spitzer et al., 2006; MBI: West; Li-Sauerwine et al., 2020; Maslach & Jackson, 1986; CD-RISC2: Vaishnavi et al., 2007) with final validation through structural equation modeling (SEM) using Stata 17.0 (StataCorp, 2022).

## Results

The study sample included 533 healthcare workers (87.2% female; mean age = 42.6 ± 20.9 years) from ambulatory (68% response rate) and rural hospital settings (39-53% response rates). Participants represented diverse roles: allied health professionals (29.3%), physicians (13.3%), nurses (8.0%), and administrative/support staff (12.3%) (Table 1:Survey Sample Characteristics).

**Table 1.** Survey Sample Characteristics (N=533)

| Variable |  | N (%) | Missing |
| --- | --- | --- | --- |
| Age (years) |  |  | 4 |
| 18 – 29 |  | 60 (11.3) |  |
| 30 – 44 |  | 218 (41.2) |  |
| 45 – 64 |  | 219 (41.4) |  |
| 65 or older |  | 22 (4.2%) |  |
| Prefer not to answer |  | 10 (1.9%) |  |
| Site |  |  | 0 |
| 1 |  | 376 (70.5) |  |
| 2 |  | 80 (15.0) |  |
| 3 |  | 77 (14.4) |  |
| Years Working in the Organization |  |  | 3 |
| < 1 |  | 97 (18.3) |  |
| 1 – 2 |  | 80 (15.1) |  |
| 3 – 5 |  | 96 (18.1) |  |
| 6 – 10 |  | 79 (14.9) |  |
| > 10 |  | 169 (31.9) |  |
| Prefer not to answer |  | 9 (1.7) |  |
| Position Within the Organization |  |  | 31 |
| Clinician/Provider |  | 281 (56.0) |  |
| Receptionist/Office Assistant |  | 118 (23.5) |  |
| Leadership/Administration |  | 62 (12.4) |  |
| Therapist/Technician |  | 25 (5.0) |  |
| Other/Non-patient contact |  | 16 (3.2) |  |

The COS demonstrated excellent internal consistency (α = 0.94) and sampling adequacy (KMO = 0.923). Principal component analysis revealed a robust three-factor structure: Component 1 (eigenvalue = 7.18, 65.3% variance), Component 2 (eigenvalue = 1.11, 10.1%), and Component 3 (eigenvalue = 1.06, 9.6%), collectively explaining 84.9% of the total variance among the items. Parallel analysis with scree plots confirmed this three-factor solution. The confirmatory factor analysis model showed excellent fit (RMSEA = 0.049, 95% CI [0.035, 0.062]; CFI = 0.992), with modifications retaining covariances between three key organizational support items. (Figure 1) The 3 subfactors are identified as Organizational Support, Access to Peer Support, and Availability of Support. (Table 2) Cronbach’s alpha for the 3 subscales were: 0.8264, 0.9187, and 0.9732, respectively.

**Table 2.** Item-Level Summary for the Newly Developed Healthcare Worker Culture of Well-being scale and Subscales: Descriptions, Endorsement Rates, and Factor Loadings from Analysis (N=533) Comparative Fit Index (CFI) and Root Mean Square Error of Approximation (RMSEA) for the Confirmatory Factor Analysis that produced these factor loadings are 0.992 and 0.049, respectively.

| Item | Description | Standardized<br>Factor Loading | Mean | SD | Missing |
| --- | --- | --- | --- | --- | --- |
|  | **Organizational Support** |  |  |  |  |
| 1 | I can count on my supervisor for support when I need it. | 0.59 | 3.33 | 0.86 | 0 |
| 2 | I trust the management at my organization. | 0.70 | 2.98 | 0.94 | 3 |
| 3 | My organization is committed to employee health and well-being. | 0.66 | 3.00 | 0.92 | 1 |
|  | **Access to Peer Support** |  |  |  |  |
| 4 | a) My organization provides a confidential peer support program for employees who have experienced a stressful work-related event. | 0.75 | 3.55 | 1.25 | 1 |
| 5 |  | 0.72 | 3.43 | 1.43 | 1 |
| 6 | b) I know who to contact for peer support in my organization if I experience a stressful work-related event. | 0.78 | 3.33 | 1.33 | 3 |
|  | c) I have prompt access to trained peer support staff who can help me if I experience a stressful work-related event. |  |  |  |  |
|  | **Availability of Support** |  |  |  |  |
| 7 | a) To listen to me without judgment. | 0.87 | 3.91 | 1.16 | 1 |
| 8 | b) To empathize with my situation. | 0.87 | 3.97 | 1.11 | 0 |
| 9 | c) To help me process my experience. | 0.91 | 3.85 | 1.18 | 2 |
| 10 | d) To provide me with emotional support. | 0.91 | 3.79 | 1.21 | 0 |
| 11 | e) To help me to be more resilient. | 0.91 | 3.72 | 1.22 | 1 |

**Figure 1.**
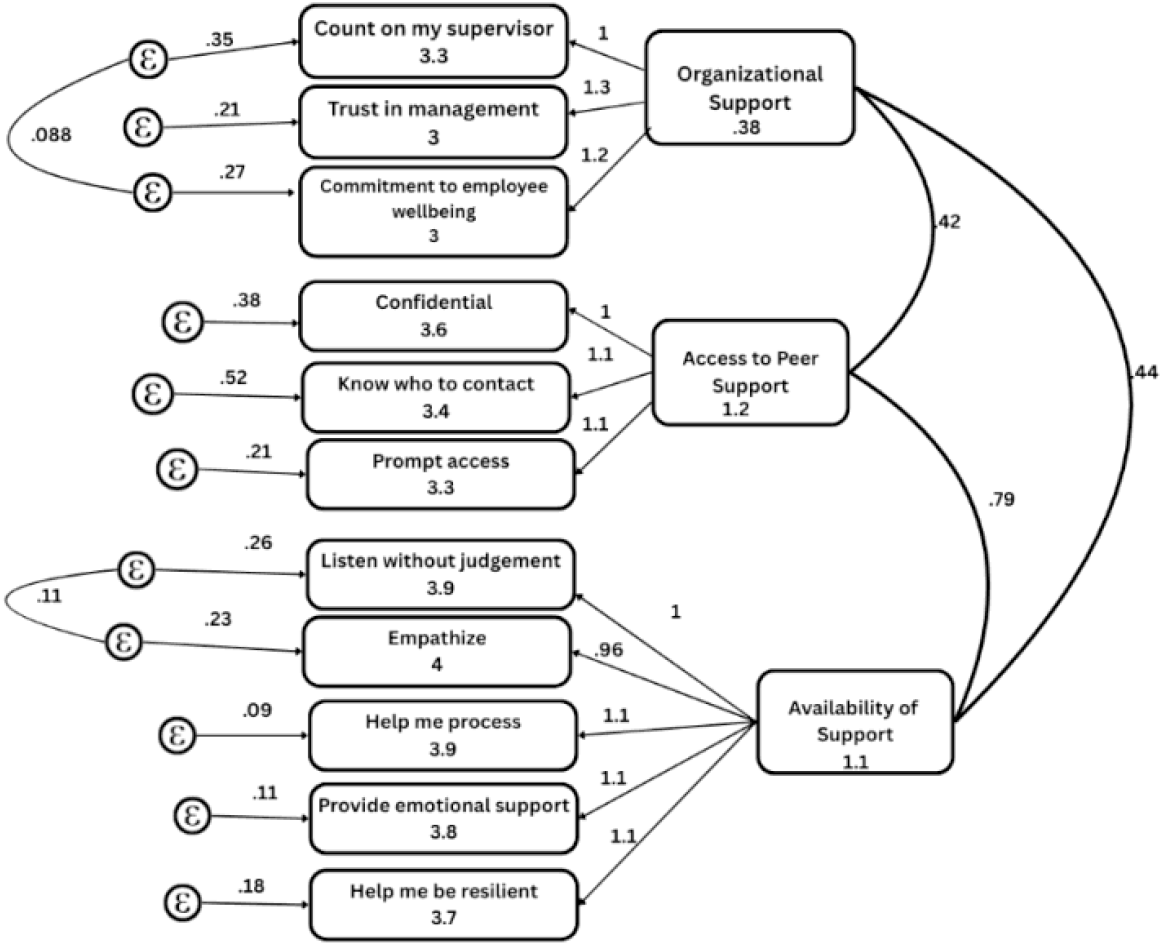
Confirmatory Factor Analysis of Healthcare Worker Culture of Well-Being (N=533) This Figure shows a structural equations modeling diagram of the confirmatory factor analysis reported in this study. Latent variables are shown in circles while observed indicators (questions from the survey) are shown in squares. Residual errors e for each indicator are shown in circles. Numbers on pathways from latent variables to indicators are unstandardized(confirm?) factor loadings. Numbers on curved double-headed arrows between latent variables show correlations among the latent variables.

We developed a support score that was an unweighted summary of the 11 item scores. As expected, the composite score had a positive correlation with resilience (r = 0.30) and negative correlation with anxiety and burnout (r = -0.35 and -0.47, respectively), indicating support systems significantly influence mental health outcomes. Canonical correlation analysis (r = 0.60) revealed that trust in management and access to confidential peer support were the strongest predictors of variance in well-being measures, while burnout showed inverse relationships with support constructs. These findings underscore how organizational trust and peer support mechanisms may buffer against burnout while enhancing resilience, highlighting their critical role in workplace well-being interventions.

## Discussion

Supporting healthcare workers experiencing distress in the workplace, including second victims, is essential to maintaining the well-being of the healthcare workforce (Seys et al., 2013; Wu, 2000). The Healthcare Worker Culture of Support Scale (COS) was developed to assess healthcare professionals’ awareness and attitudes about the availability of support resources, and the organizational culture that underpins well-being. To our knowledge, this is the first tool to measure awareness of organizational support programs for HCWs. The new survey differs from Second Victim Experience and Support Tool (SVEST), which focuses on the psychological impact of second victim experiences and the quality of post-event support (Burlison et al., 2017). The new survey also differs from the WITHSTAND-PSY survey which measures a second victim’s emotional distress before and after a patient adverse event. The COS evaluates HCWs’ awareness, attitudes and perceived accessibility of support resources within their institution regardless of prior exposure to adverse events. Unlike the WITHSTAND-PSY, the COS measures HCWs’ perceptions of organizational support, access to formal peer support, and availability of peer support within their workplace settings.

We found that the COS demonstrates a clear three-factor structure: 1) Organizational Support, 2) Access to Peer Support and 3) Availability of Support. Together, these three factors account for 84.9% of the variance in the data. Confirmatory SEM, adjusted for key covariances (items 9a–9b and 4–7), yielded excellent fit (RMSEA = 0.049, CFI = 0.992), and internal consistency was very high (Cronbach’s α = 0.94). The internal consistency coefficients were 0.82 or higher for the three subscales. The psychometric properties of the instrument were also supported by using PCA for dimensional reduction, CCA to assess associations between support constructs and mental health variables, and SEM for validation. The findings suggest that the COS and the three subscales could be useful for assessing the supportiveness of the work environment.

The three dimensions of COS illustrate different aspects of an organization’s culture of support. Organizational Support encompasses worker beliefs and the commitment of leaders that make help-seeking a normalized behavior. Items like “I trust the leadership in my organization” and “My organization is committed to the health and well-being of its staff” load most heavily here, reflecting a belief in the organization’s commitment to the well-being of the healthcare workforce. Access to Support describes access to concrete resources, from knowledge of who to contact to find peer support to access to trained staff when needed. The emergence of this as a separate factor highlights that knowing the logistical pathways to utilize support is critical. Availability of Support describes individual beliefs about stigma, the value of empathetic listening, and resilience building. Statements like “To listen to me without judgement” and “To help me to build more resilience” demonstrate how perceptions influence engagement with support programs. The combination of these constructs offers a comprehensive, proactive view of how culture, resources, and attitudes intersect to influence second-victim support.

The three-factor structure of this scale aligns with established occupational health frameworks. Within the Job Demands–Resources model, organizational support reflects institutional resources that buffer job stressors; peer support functions as a key social resource that enhances coping and engagement; and availability of support captures motivational resources that reduce stigma and encourage help-seeking. (Demerouti, et al., 2001) From a socio-ecological perspective, these dimensions operate across multiple levels: organizational (leadership and policies), interpersonal (peer relationships), and individual (perceptions of accessibility and stigma). (Habeger et al., 2022). Together, they underscore that HCW well-being is shaped by the interaction of institutional structures, social networks, and individual beliefs.

COS showed good construct and criterion validity. Higher survey scores were positively associated with better mental health profiles: greater Organizational Support was linked to lower anxiety, lower burnout, and higher resilience; Access to Peer Support was tied to lower burnout and showed the strongest relationship with individual resilience, followed closely by Availability of Support. Collectively, these patterns suggest that the scale not only reflects current well-being but may predict key outcomes such as staff retention and on-the-job performance. Organizations can use this survey to assess healthcare workers’ perceptions of the workplace culture of support and to inform the development of interventions that foster a more supportive work environment. By collecting the survey at intervals and linking scores to metrics such as retention, incidents, and employee engagement, institutions can iteratively monitor cultural change and conduct more systematic evaluations of their programs.

Although this was developed and validated among rural healthcare workers and in ambulatory practices, its underlying constructs are broadly relevant to healthcare delivery in diverse settings. Rural healthcare settings provide a unique testing ground due to resource limitations, workforce shortages, and professional isolation, which magnify the importance of organizational culture and peer support. However, these same dimensions are present in urban academic centers, large health systems, and international contexts, albeit expressed differently. The psychometric strength and theoretical grounding of this tool suggest that it can be adapted and applied across healthcare environments.

Despite its strong psychometric profile and clear three-factor structure, the COS scale has areas that warrant further attention. The high inter-item correlation between culture and access items suggests potential redundancy, indicating an opportunity to streamline or collapse overlapping questions to sharpen the survey’s discriminatory power. Likewise, items tied to stigma perceptions and help-seeking norms could be targeted in pilot interventions and are expected to show the earliest, most pronounced score shifts when leadership training or resource-visibility campaigns are introduced. Finally, while our sample size of 533 participants offered robust SEM validation and links to mental health outcomes, broader testing across varied healthcare settings is needed to confirm the scale’s responsiveness to organizational change and its predictive value for performance outcomes.

The study had some limitations. The scale was tested on a limited number of rural and ambulatory sites in the Mid-Atlantic. Variable response rates across sites raises the possibility of selection bias. These factors may affect its generalizability to larger or more diverse healthcare systems. However, there is an opportunity for future research to further validate and replicate the scale’s factor structure. Another potential limitation was the reliance on self-report measures and potential reporting and social desirability bias, which could have increased the internal consistency of the subscales. As noted above, the high internal consistency reliability coefficient (Cronbach’s α = 0.92) may reflect excessive redundancy among items and need for further refinement. However, the internal consistency coefficients were lower for the three component subscales. This reduces the need for further item reduction.

## Conclusion

In summary, preliminary analyses of the Healthcare Worker Culture of Support scale suggests that it provides a reliable and valid measurement of HCW’s knowledge, attitudes and access to supportive resources. With further refinement and validation, this overall scale and each of the subscales could be useful for healthcare organizations aiming to improve the institutional culture of support, resilience, and well-being for members of their workforce.

## Data Availability

All data produced in the present study are available upon reasonable request to the authors

## Acknowledgments

We would like to acknowledge the contributions of all of the participating healthcare workers in completing the survey.

The Health Resources and Services Administration (HRSA), U.S. Department of Health and Human Services (HHS) provided financial support for this publication. The award provided 100% of total costs and totaled $2,303,362. The contents are those of the authors. The information therein may not reflect the policies of HRSA, HHS, or the U.S. Government.

